# Comparing AI and Human Coding of NIH Grant Abstracts to Identify Innovations in Opioid Addiction Treatment

**DOI:** 10.64898/2026.02.13.26346235

**Authors:** Sarah A. Alkhatib, Nadiya Jiwa, Dallin Judd, Justin M. Luningham, Ginnie Sawyer-Morris, Merve Ulukaya, Todd Molfenter, Faye S. Taxman, Scott T. Walters

## Abstract

Large language models (LLMs) are increasingly used for qualitative analysis in substance use research, yet their performance relative to human coders remains underexplored. This study compares ChatGPT-4.0 with human coders in identifying and describing the core innovation of NIH grants focused on reducing opioid overdose. A total of 118 NIH HEAL Initiative grant abstracts were independently coded by ChatGPT and humans to generate innovation descriptions, which were then evaluated by both human raters and ChatGPT for depth/detail and relevance/completeness using 5-point Likert scales. Identical instructions were used across all coding and evaluation stages. ChatGPT-generated descriptions were consistently rated higher than human-generated descriptions on both dimensions. Human evaluators rated ChatGPT outputs at an average of 4.47 for both depth/detail and relevance/completeness, compared to 3.33 and 3.24 for human outputs, respectively (F(1,176)=133.9, p<0.001). These findings suggest that LLMs, when carefully prompted, can enhance the efficiency and quality of qualitative research evaluation.

## Introduction

### Advances in Qualitative Coding

Systematic reviews provide the critical syntheses of studies that can inform scientific, clinical, and policy decisions (1). Traditionally, systematic reviews rely on time-consuming and intensive manual methods to code qualitative data, which can limit the scope and speed of analysis. In qualitative research, ‘coding’ means systematically labeling and categorizing text to capture key themes or constructs. This process typically starts with familiarizing oneself with the data, followed by developing a coding scheme, applying these codes to the data, and analyzing the coded data to draw conclusions (2). Given the complexity and volume of new research, there is growing interest in leveraging artificial intelligence (AI) to streamline and enhance the coding process (3, 4). However, questions remain about the reliability, validity, and sensitivity of AI-generated coding compared to manual methods. This study examines the comparability of AI and manual coding in identifying the basic “innovation” of NIH-funded studies to reduce opioid overdoses. Innovation coding requires a relatively high level of precision and contextual understanding, making it an excellent domain for evaluating the capabilities of AI in qualitative analysis.

Programs like ChatGPT can enhance the efficiency of qualitative coding by rapidly processing datasets and providing initial thematic analyses that human researchers can further refine (5). This approach not only saves time but also allows for more consistent and comprehensive coverage of the data (6, 7). For instance, a study by Lennon et al. (2021) found that AI-assisted coding achieved a level of thematic depth comparable to human coding (8). Another study showed that AI tools allowed for the identification of emergent themes that might be overlooked by manual coders due to cognitive biases (9). At the same time, it is essential to address AI’s tendency to favor its own outputs compared to human coding. For instance, research has shown that while AI-generated thematic codes can be detailed and expansive, their validity and reliability still requires human cross-validation to ensure accuracy (10).

In sum, despite the increasing interest in using AI for qualitative research, there exists a noticeable gap in understanding how AI performs against manual coding (4). Authors have discussed the potential of AI to enhance research efficiency, but they also note challenges such as the need for validation and the risk of AI introducing bias into qualitative analysis (4). This gap is particularly urgent for assessing the innovation of research projects in areas that are rapidly evolving (10).

In this study, we focused on the “innovation” described in NIH-funded studies to reduce opioid overdoses. Understanding the innovation being tested in a study can help to better gauge the breadth of a funding portfolio, connect researchers with others who are testing similar innovations, and identify the ideas and settings that have been most effective at mitigating opioid-related harms (and by extension those that are being missed).

### Innovations in Overdose Prevention

Over the last decade, the US has seen a sharp rise in overdose deaths, which totaled approximately 90,000 in the 12 months preceding July 2024 (11). In response, the National Institute of Health (NIH) launched the Helping to End Addiction Long-term (HEAL) initiative to “speed scientific solutions to stem the national opioid public health crisis (12). Between 2018 and 2022, the HEAL initiative funded an estimated 923 unique research studies (i.e., excluding grant renewals and supplements) through more than 200 funding mechanisms. The HEAL portfolio includes a broad range of studies from basic science to community implementation. For instance, early-stage research (T0-T1) examines basic science or proof of concepts that lead to later-stage research (T2-4) focusing on translation to clinical or relevant practice.

One way to understand the breadth of this research portfolio is in terms of the specific innovation, idea or practice being tested in a study (13). A previous study conducted by this group used a manual coding process based on the Consolidated Framework for Implementation Research (CFIR) to extract relevant innovation characteristics from the full body of HEAL-funded projects (12). Coders used grant abstracts available in NIH RePORTER. The result was a high-level overview of the portfolio, including areas of strength and suggestions for future investment. For instance, the analysis concluded that (as expected) most HEAL-funded studies either addressed pain (37.4%) or substance use (46.3%). T0-T1 studies tended to be conducted in the laboratory (35.4%), while T2-T4 studies were primarily conducted in hospital settings (21.6%), with relatively little representation in community settings (12.9%). When considering the innovation target, 88% mentioned gaps in existing services, 80% mentioned human behavior and problems (e.g., overdose), and 41% discussed existing gaps in research (12). Based on the analysis, the study recommended better prioritizing underserved populations, engaging community stakeholders, and translating evidence-based practices into non-clinical settings.

The current analysis focuses on a subgroup of 125 HEAL-funded studies from the “Translation of Research to Practice for Treatment of Opioid Addiction” (12) subcategory. This subgroup (referred to as the “Translation” portfolio) was concerned with testing the integration and implementation of interventions in community settings (all T2-4). Examples include tests of new medications to reduce opioid use disorder in hospital settings, harm reduction approaches to reduce drug-related consequences in the community, and behavioral interventions to reduce overdoses after discharge from jails.

### The CFIR Coding Process

In the study described above, a data collection tool was developed using CFIR domains (e.g., outer setting, inner setting), coding guidelines, and the relevant literature (2, 13, 14). A research team (n = 8) was trained to code projects using the CFIR tool via three information sessions, practice case assignments, and an evaluation session. Researchers began coding in a Qualtrics form after they attained 90% consensus with the trainers (n = 2). Once coding was underway, trainers held monthly progress meetings to share updates, obtain feedback about the tool, and address emerging questions. Between June 2023 and October 2024, the team coded a total of 923 HEAL abstracts using a 54-item CFIR data collection tool. Once coding was complete, data were exported from Qualtrics to Stata 18.0.

### Rationale for the Present Study

In this project, we drew on a subset of the previously coded HEAL portfolio and applied the same coding framework. Whereas the original study relied exclusively on human hand-coding, our goal was to test whether AI could perform the same tasks and how its outputs compared in quality to those of human coders in the previous study. Thus, this study aims to answer the following questions: 1) How comparable are innovation descriptions as determined by ChatGPT and human coders? and 2) What is the quality of those innovation descriptions as judged by humans and ChatGPT?

We used two metrics to assess the quality of the innovation descriptions: depth/detail (D&D) and relevance/completeness (R&C). A similar methodology was employed by Morgan et al. (2023) to evaluate AI’s performance in qualitative coding within healthcare research (15). Their study compared AI-generated codes with those produced by human analysts, focusing on thematic accuracy, coding consistency, and the granularity of insights. The findings indicated that while AI was proficient in identifying broad themes, it often lacked the nuanced understanding that human coders provided, particularly in areas that required deep contextual interpretation.

## Methods

### Sample and Procedures

As previously mentioned, we limited our analysis to grants available in the NIH RePORTER database that HEAL classified as “Translation of Research to Practice for Treatment of Opioid Addiction” (n=125). We further limited the scope to projects funded by an “R” mechanism (e.g., R01, R03, R21, R34), as these were most likely to represent discrete research ideas (n=45). This eliminated other kinds of grants whose primary purpose was to train or support, for instance, training programs and coordinating centers.

Human coders in this study were trained using the same procedures described by Sawyer-Morris et al. (2025), including three training sessions, practice case assignments, and a consensus check requiring at least 90% agreement before independent coding. This ensured consistency with the previous portfolio analysis and allowed us to directly compare human and AI performance on the same coding task.

The task was structured into two main stages. In the first stage, human coders and ChatGPT were asked the same question for each grant: “What is the innovation being tested in this study?” “Innovation” was defined as the distinctive idea, intervention, or approach that the grant proposed to explore or test. This could include novel methodologies, technologies, theoretical frameworks, or applications that differentiate it from previous studies. The human coders had no prior contextual knowledge of the projects beyond what was presented in the grant abstracts, ensuring that their coding decisions were solely based on the provided text. This was done to ensure parity with ChatGPT, which also received only the grant abstract. All coders had sufficient disciplinary background to interpret NIH abstracts. The abstracts were similarly inputted into ChatGPT v4.0, replicating the instructions given to human coders. A standardized statement was prompted to ChatGPT: “I will provide you with an abstract and would like your assistance in answering the following 4 questions related to it.” This statement primed ChatGPT for a task-oriented response to ensure alignment with human coding processes. Both ChatGPT and human coders were given the same four guiding questions: (1) What primary innovation or advancement is being pursued or tested in this research project?; (2) What type of innovation does this research primarily fall under? List all category numbers that correspond to the type of innovation?; (3) Select the science type(s) that best describe the nature of the study being conducted. List all category numbers that correspond to the type of science.; and (4) Briefly summarize the primary outcome this research aims to investigate or achieve. For the purposes of this paper, we focused exclusively on Question 1, as our primary interest was the innovation of the grant.

In the second stage, once the innovation descriptions were created, 4 human coders and ChatGPT gave two ratings per abstract: one for depth/detail and one for relevance/completeness on a Likert scale of 1-5. The four questions served as prompts to generate the descriptions, which were then rated using these two criteria (depth/detail and relevance/completeness). The same set of human coders also served as raters, but all assignments were randomized and anonymized so that coders did not know which outputs they were evaluating.

1. Depth/Detail (D&D): The richness and specificity of the description, with higher scores indicating greater granularity in capturing detail and nuance.
2. Relevance/Completeness (R&C): The degree to which the description aligned with the core elements of the project and fully captured the intended innovation.

We used Likert scales [1–5] for both metrics, with higher scores indicating better performance on that metric. For Relevance & Completeness (R&C), innovation descriptions were rated on their ability to capture all relevant aspects of the innovation while avoiding irrelevant or extraneous information. A score of 1 indicated a response that was off-topic or failed to encompass the essential components of the innovation, while a score of 5 represented a highly relevant and complete response to the key aspects of the research’s novelty. These Likert scale ratings enabled a structured comparison of the innovation descriptions, helping to quantify the strengths and weaknesses of ChatGPT’s and human coders’ outputs in terms of both depth of analysis and relevance to the task at hand.

To gauge the consistency of ChatGPT’s innovation descriptions, 15 grant abstracts were independently entered into ChatGPT by four coders on separate accounts using identical instructions. Responses showed high consistency, with ≥90% semantic overlap in identified innovations and no substantive contradictions. This confirmed that results were not dependent on a single session or login. Finally, we were curious whether the length of responses influenced the quality of ratings, so we conducted a correlation analysis between the character count of the innovation descriptions and their D&D and R&C scores using all original 125 abstracts. We also assessed inter-rater reliability among human coders using intraclass correlation coefficients (ICCs) for both D&D and R&C ratings.

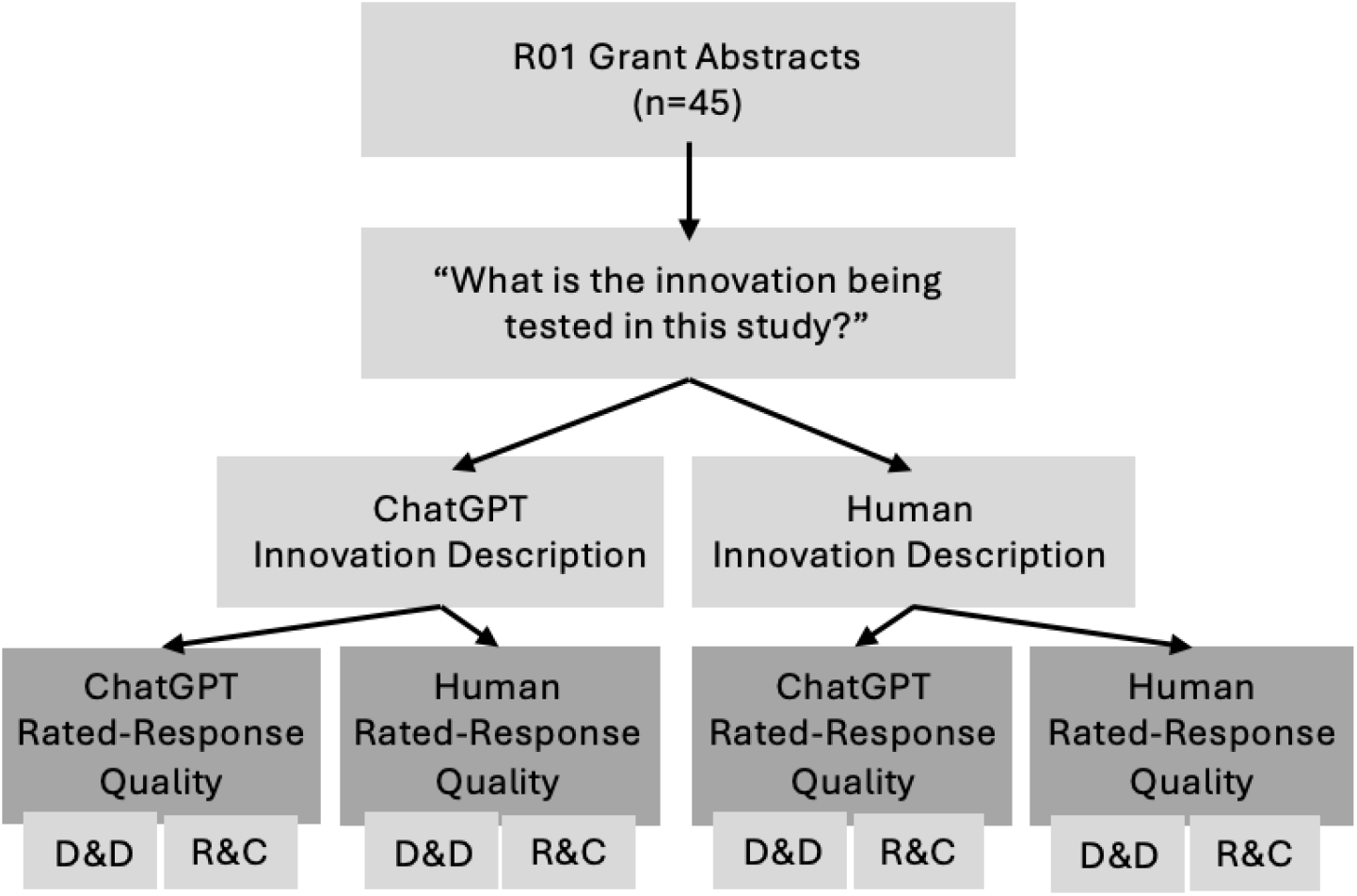

### Analysis Plan

To determine differences between human and ChatGPT innovation responses, a two-way analysis of variance (ANOVA) was conducted for both D&D and R&C scores. The ANOVA included the following factors: 1) Output Source: Human-generated descriptions versus ChatGPT-generated descriptions. 2) Rater Source: Human raters versus ChatGPT rating the outputs. This factorial design allowed us to test: The main effect of output source (human vs. ChatGPT-generated outputs), the main effect of the scorer source (human vs. ChatGPT as raters), and the interaction effect between the output source and scorer source (i.e., whether the relative differences in scores assigned by human and ChatGPT scorers depend on whether the output was generated by humans or ChatGPT).

The ANOVA was conducted separately for D&D and R&C metrics to determine whether the observed differences in mean scores across groups were statistically significant. Assumptions of normality of residuals and homogeneity of variance were tested using Shapiro–Wilk and Levene’s tests, respectively. Both assumptions were met for D&D and R&C outcomes, supporting the use of ANOVA. Post-hoc descriptive analysis was also performed to examine patterns in mean ratings, providing additional context for the statistical results.

## Results

The comparison of human and ChatGPT 4.0 is shown in Table 1. There were significant differences in both D&D and R&C ratings. In terms of D&D, there was a significant main effect of output source, (F(1, 176) = 141.19, P < 0.001), indicating that both humans and ChatGPT consistently rated ChatGPT output more favorably compared to human outputs, with average ratings of 4.23 and 2.97, respectively. Interestingly, human coders tended to assign more favorable ratings regardless of the output source (3.9 average rating assigned by humans, compared to 3.3 by ChatGPT), F(1, 176) = 31.68, P < 0.001). The interaction effect between the output source and scorer source was not significant (F(1, 176) = 1.56, P = 0.21).

**Table 1:** Overall Averages of Depth & Detail (D&D) and Relevance & Completeness (R&C) Scores *(Means and Standard Deviations)*

|  | D&D |  |  | R&C |  |  |
| --- | --- | --- | --- | --- | --- | --- |
|  | Human Rater | GPT Rater | Output Average | Human Rater | GPT Rater | Output Average |
| Human Output | 3.33<br>(1.00) | 2.60<br>(0.69) | 2.97<br>(0.93) | 3.24<br>(1.00) | 2.89<br>(0.78) | 3.07<br>(0.91) |
| GPT Output | 4.47<br>(0.66) | 4.00<br>(0.37) | 4.23<br>(0.58) | 4.47<br>(0.66) | 4.24<br>(0.43) | 4.36<br>(0.57) |
| Scorer average | 3.90<br>(1.02) | 3.30<br>(0.89) |  | 3.85<br>(1.04) | 3.57<br>(0.92) |  |

**Table 2:** D&D ANOVA Scores.

|  | SS | Degrees of Freedom | MS | F-value | P-value |
| --- | --- | --- | --- | --- | --- |
| Output Source | 72.2 | 1 | 72.2 | 141.19 | < 0.001 |
| Scorer Source | 16.2 | 1 | 16.2 | 31.68 | < 0.001 |
| Output x Scorer Interaction | 0.8 | 1 | 0.8 | 1.56 | 0.21 |
| Error | 90 | 176 | 0.51 |  |  |
| Total | 179.2 | 179 |  |  |  |

**Table 3:**
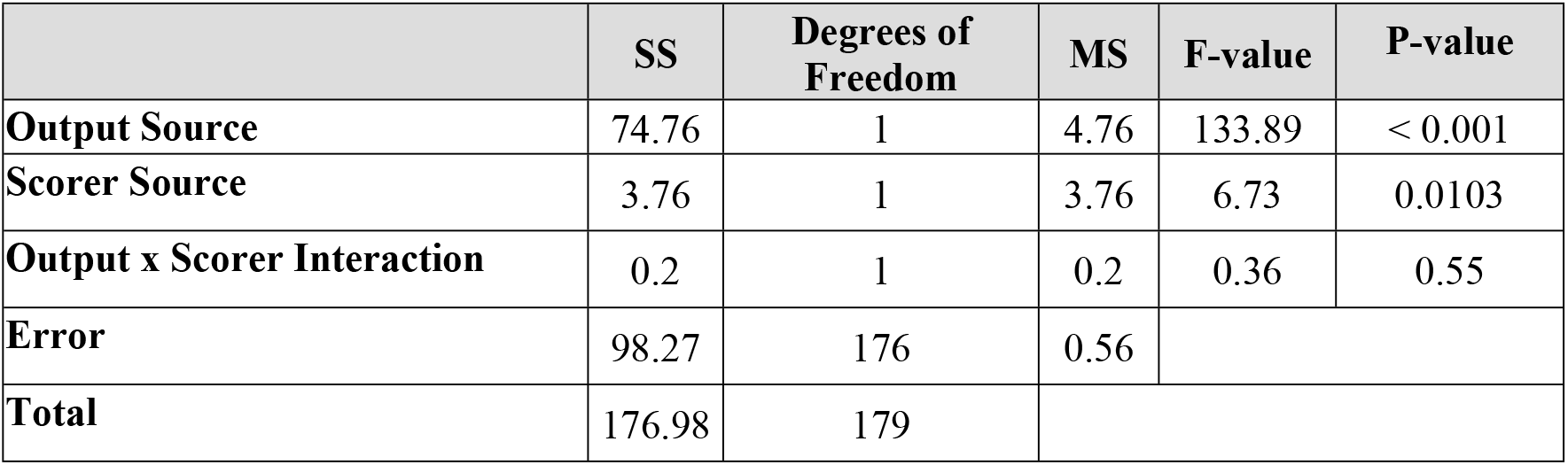
R&C ANOVA Scores.

A similar pattern was detected for R&C ratings. ChatGPT outputs tended to be rated more highly than human outputs, with an average rating of 4.36 and 3.07, respectively, F(1, 176) = 133.89, P < 0.001). The scorer source was also statistically significant (F(1, 176) = 6.73, P = 0.0103). Humans tended to score outputs more favorably, compared to ChatGPT (M = 3.85 and 3.57, for humans and ChatGPT, respectively).

As shown in Table 1, the ChatGPT-generated outputs were rated higher than human-generated outputs on both Depth & Detail (D&D) and Relevance & Completeness (R&C). The average D&D score for ChatGPT outputs was 4.23 (SD = 0.58), significantly exceeding the average score for human outputs (M = 2.97, SD = 0.93). Similarly, ChatGPT outputs achieved a higher average R&C score of 4.36 (SD = 0.57) compared to 3.07 (SD = 0.91) for human outputs. Scorer ratings revealed a consistent pattern: human scorers assigned higher average scores overall (D&D: M = 3.90, SD = 1.02; R&C: M = 3.85, SD = 1.04) compared to GPT scorers (D&D: M = 3.30, SD = 0.89; R&C: M = 3.57, SD = 0.92). ANOVA results confirmed significant main effects for both metrics. For D&D, there was a significant main effect of output source, F(1,176) = 141.19, p<0.001, indicating higher ratings for ChatGPT outputs. A significant main effect of scorer source was also observed, F(1,176) = 31.68, p<0.001, with human scorers assigning higher ratings than GPT scorers. However, the interaction between output source and scorer source was not significant, F(1,176) = 1.56, p=0.21. A similar trend was observed for R&C scores. The main effect of output source was significant, F(1,176) = 133.89, p<0.001, favoring ChatGPT outputs. The scorer source effect was also significant, F(1,176) = 6.73, p=0.0103, showing a tendency for higher ratings from human scorers. No significant interaction effect was found, F(1,176) = 0.36, p=0.55, indicating consistent differences across scorer types.

To assess whether the length of innovation descriptions influenced their ratings, we conducted a correlation analysis between the character count of descriptions and their D&D and R&C scores. The analysis revealed a positive modest correlation between response length and both D&D (r = 0.34, P < 0.01) and R&C (r = 0.29, P < 0.01) scores.

## Discussion

This study evaluated the performance of ChatGPT 4.0 vs. manual coding in assessing the innovation in NIH grant abstracts related to opioid overdose. Our analysis revealed three key findings: 1) ChatGPT-generated outputs were consistently rated higher in terms of depth/detail and relevance/completeness than human-generated outputs; 2) manual coders tended to assign higher ratings than ChatGPT scorers for both human and ChatGPT-generated outputs; and (3) response length showed a modest but positive correlation with the quality of the ratings. These findings have important implications for the integration of AI tools like ChatGPT in qualitative research and evaluation processes.

### Superior Quality of ChatGPT Outputs

The most striking finding from this study is that ChatGPT-generated innovation descriptions outperformed those of humans across both D&D and R&C metrics. ChatGPT’s average D&D score of 4.23 compared to the human average of 2.97, and R&C scores of 4.36 versus 3.07, suggest that ChatGPT’s responses were judged to be both more detailed and more relevant. This aligns with the growing body of evidence that large language models (LLMs) can process and synthesize complex textual information at a level of detail that may surpass that of human coders, particularly in tasks requiring structured analysis of large datasets (16-18). The superior ratings for ChatGPT outputs suggest that AI-driven coding could offer significant advantages in terms of efficiency and richness of generated insights. These findings highlight the potential for LLMs to complement or even enhance traditional qualitative analysis workflows. While human expertise remains critical for nuanced interpretation, particularly in highly subjective or context-dependent tasks, AI tools like ChatGPT can augment human efforts by providing comprehensive, high-quality descriptions that meet standardized evaluation criteria. This is particularly relevant for large-scale content analysis, where AI could reduce the time and labor required while maintaining or improving the depth of insights generated.

### Humans Rate More Favorably

A second finding was that human coders tended to assign more favorable ratings overall compared to ChatGPT, regardless of whether they were evaluating human or ChatGPT descriptions. This discrepancy could reflect differences in how the two scoring systems interpret the evaluation criteria. For instance, human raters might value more subtle elements of writing that align with human cognition and communication, whereas ChatGPT may use more rigid or algorithmic interpretations of relevance and detail. This tendency of human scorers to rate both sets of outputs higher also raises questions about potential biases in human evaluation. It may suggest that humans, when assessing outputs from an LLM, are inclined to overestimate the value of responses due to the novelty or perceived authority of AI outputs. Alternatively, it could suggest that human scorers, familiar with qualitative coding processes, may have been more lenient in their evaluations than the ChatGPT system, which may have adhered more strictly to the predefined criteria. Further research is needed to explore these differences and to better understand how human and AI evaluators differ in their application of qualitative metrics

### Response Length Modestly Correlated with Quality

The correlation between response length and quality ratings for both D&D (r = 0.34) and R&C (r = 0.29) suggests that while longer responses were more likely to be rated as higher quality, length was not solely responsible for the superior performance of ChatGPT. This finding is critical as it underscores that high-quality coding depends not just on word count but on the substance and coherence of the content. This suggests that AI tools like ChatGPT can generate concise yet high-quality analyses when appropriately instructed, which has implications for optimizing AI-generated outputs in research contexts. Future work should focus on refining prompt structures and instructional guidelines for LLMs to ensure that response length is balanced with relevance and detail, maximizing the effectiveness of AI in qualitative research.

### Limitations

Our study has several limitations that may impact the generalizability of the findings. First, we focused on two specific evaluation criteria—D&D and R&C—which may not fully capture the range of quantitative coding tasks, particularly those requiring nuance or contextual understanding. Additionally, we did not explore the decision-making processes of human coders or the AI, leaving questions about how they arrive at their conclusions. Although it would have been feasible to request short justifications from human coders or to prompt ChatGPT to provide reasoning, we did not do so in this study in order to keep coder burden minimal and maintain parity between human and AI outputs. Future work should incorporate reasoning or ‘explanation’ prompts to enable a richer comparison of not only the outputs but also the underlying decision-making processes. The study’s reliance on a single AI model, ChatGPT 4.0, limits the applicability of our findings to other models, particularly as AI technology evolves rapidly. Finally, AI performance is highly dependent on prompt design, and variations in prompts could yield different outcomes, underscoring the need for further research into prompt standardization and cross-model comparisons.

### Strengths and Future Recommendations

The findings from this study demonstrate that AI methods, particularly ChatGPT 4.0, can reliably perform coding tasks at a level comparable to or exceeding human coders in many aspects. The potential to integrate LLMs into research evaluation processes offers substantial benefits in terms of efficiency, scalability, and consistency. However, it is important to consider the limitations of AI-driven analysis, particularly in tasks requiring high levels of contextual interpretation, where human judgment remains indispensable.

Future research should build on these strengths by incorporating additional evaluation metrics, such as clarity, originality, and grammatical correctness, to provide a more comprehensive assessment of response quality. Expanding the analysis to diverse datasets and task conditions will help validate the generalizability of our findings across different domains. Furthermore, integrating qualitative methods, such as coder interviews or think-aloud protocols, could offer deeper insights into the decision-making processes behind evaluations. Comparative studies involving multiple AI models would also be beneficial in identifying the specific strengths and limitations of various AI technologies.

Our findings raise important considerations for how large language models might be incorporated into grant review and research evaluation processes. While AI showed strong performance in generating detailed and relevant innovation descriptions, it is essential that grant applicants be assured that their work is assessed by human reviewers. The use of AI in coding or summarizing proposals should therefore be viewed as an aid to, rather than a replacement for, human judgment. Applicants and stakeholders must also be confident that AI-supported evaluations are unbiased and that the criteria used are consistent and clearly documented. Safeguards such as standardized prompts, transparency about when and how AI is used, and the continued involvement of human reviewers are likely to be necessary if AI-generated coding is to play a role in shaping funding priorities. Finally, ongoing evaluation of AI’s performance considering its rapid advancements will be essential to ensure that our understanding of its capabilities remains current and relevant.

## Conclusion

This study demonstrates the potential of ChatGPT 4.0 to enhance qualitative coding, as ChatGPT-generated outputs outperformed human coders in terms of depth and relevance when analyzing innovation content in NIH-funded projects to reduce opioid overdoses. AI showed greater efficiency and consistency in structured data environments, though human scorers tended to rate both human and AI outputs more generously. Despite these promising results, the study’s focus on structured data and a single AI model limits the generalizability of the findings. Future research should explore AI’s performance in more complex qualitative tasks and across different AI models. Overall, this study highlights the value of integrating AI into qualitative research to improve coding efficiency and quality.

## Data Availability

All relevant data are within the manuscript and its Supporting Information files.

## Acknowledgements

An acknowledgement to all authors for their contributions to the study.

## Acknowledgements

The authors have no conflicts of interest to disclose. This research was supported by the NIH HEAL Initiative under grant number OT2OD034479. The content is solely the responsibility of the authors and does not necessarily represent the official views of the National Institutes of Health.

## Abbreviations

AI: artificial intelligence
ANOVA: analysis of variance
CFIR: Consolidated Framework for Implementation Research
D&D: depth/detail
HEAL: Helping to End Addiction Long-term
LLM: large language model
NIH: National Institutes of Health
R&C: relevance/completeness

